# Multimodal Stress Testing and Morphologic Predictors of Ischemia in Anomalous Aortic Origin of a Coronary Artery

**DOI:** 10.64898/2026.03.12.26348294

**Authors:** Michael Jiang, Olivia McCloskey, Samantha Xu, Meghana Iyer, Tara Karamlou, Eugene H Blackstone, Elizabeth V Saarel, Austin Firth, Jeevanantham Rajeswaran, Hani K Najm, Gosta B Pettersson, Shinya Unai, Joanna Ghobrial, Cleveland Clinic Adult Anomalous Aortic Coronary Origin of the Aorta Working Group

## Abstract

**Background:** Anomalous aortic origin of a coronary artery (AAOCA) can cause myocardial ischemia and sudden cardiac death. The optimal stress-testing strategy and impact of coronary morphology on ischemia remain unclear. We assessed the effect of coronary morphology on stress-test completion and results across multiple test modalities.

**Methods:** This retrospective cohort study included 531 adults with AAOCA at our institution (7/2015 – 3/2023). Coronary morphology was characterized by the anomalous coronary (right [RCA], left main [LMCA], left anterior descending [LAD], left circumflex) and the course type (intramural, interarterial-only, transseptal, and other [prepulmonic and retroaortic]). Exercise and pharmacologic stress tests were positive if ischemia included the territory of the anomalous coronary. A mixed-effect logistic regression modeled the odds of a positive test based on morphology, comorbidities, and modality. A random forest regression analyzed the stress iFR as a continuous outcome.

**Results:** Stress test results were available for 396 (75%) of patients (age 50 ± 17 years; 42% female). Stress testing included 699 ECGs, 198 echocardiograms, 288 SPECTs, 133 PETs, and 103 dobutamine iFR studies. Completion of invasive dobutamine iFR (versus noninvasive-only) stress testing was associated with high-risk coronary morphology, p<0.001. Coronary morphology that trended toward higher adjusted odds of ischemia included the anomalous LMCA (OR: 2.1, p=0.054) and intramural course (OR: 1.9, p=0.14). Compared to ECG, iFR had higher adjusted odds of a positive result (OR: 27, p<0.001), followed by PET (OR: 9.0, p<0.001). In the random forest regression, stress iFR value was lowest for LAD (0.75) compared to LMCA (0.83) and RCA (0.84). For course type, transseptal (strongly correlated with the anomalous LAD) had the lowest stress iFR (0.77), followed by intramural (0.83), and interarterial (0.88).

**Conclusions:** In our adult AAOCA cohort, high-risk coronary morphology demonstrated a borderline association with ischemia on stress testing, whereas stress test modality was the strongest determinant of ischemia detection. Invasive stress testing was reserved for higher-risk coronary morphology. These findings underscore that effective risk stratification in AAOCA integrates clinical symptoms, coronary morphology, and stress test modality. Long-term follow-up is needed to determine the optimal strategy for ischemia evaluation.

**Clinical Perspectives:** *What is new?:*

- In this single-center registry of adults with anomalous aortic origin of a coronary artery (AAOCA), 75% of patients had stress testing, enabling the largest analysis of how anomalous coronary morphology impacts stress testing practices and the presence of ischemia.
- Consistent with published data in younger AAOCA cohorts, our adult population (mean age >50 years) trended toward increased risk of ischemia with an anomalous left coronary and intramural course.
- Comparing various stress test modalities for AAOCA, instantaneous wave-free, followed by positron emission tomography and single-photon emission computed tomography, have higher odds of being positive for ischemia than electrocardiogram and echocardiograms.

*What are the clinical implications?:*

- Adults with AAOCA remain at risk for ischemia and require careful risk stratification, with coronary morphology and clinical symptoms informing stress test selection and result interpretation.
- For higher risk morphologic variants of AAOCA, consider further risk stratification with invasive coronary provocative studies to detect inducible ischemia.

## Introduction

Anomalous aortic origin of a coronary artery (AAOCA) is a congenital heart malformation and a cause of ischemia and sudden cardiac death.^1–3^ Present in approximately 0.4-0.8% of the general population, AAOCA is defined as one of the major coronary arteries arising from the aorta somewhere other than the usual sinus of Valsalva.^4^ The anomalous coronary may then course between the great arteries (interarterial), through the ventricular septum (transseptal), posterior to the aorta (retroaortic), or anterior to the pulmonary artery (prepulmonic). The interarterial course is commonly accompanied by an intramural course, in which the anomalous coronary artery ostium travels circumferentially through the aortic wall.^5^ The interarterial, intramural, and transseptal courses can exhibit baseline luminal compression exacerbated by exercise, leading to ischemia.^1,6,7^

While these morphologic factors are associated with sudden death and ischemia in children and young adults,^2,6^ their prognostic significance in older adults is less apparent.^8–11^ Acknowledging the limited evidence, guidelines for surgical repair rely on r isk stratification based on morphology, symptoms, and inducible ischemia.^12,13^ However, symptoms such as chest pain and exertional dyspnea can be nonspecific.^9,14^ Furthermore, cardiovascular comorbidities, such as coronary artery disease (CAD) in older adults, can obscure the exact cause of symptoms.^15,16^ Therefore, exercise and pharmacologic stress tests are commonly used to detect evidence for ischemia and risk-stratify patients. Common functional exercise or pharmacologic stress testing modalities for AAOCA include electrocardiography (ECG), echocardiography, single-photon emission computed tomography (SPECT), positron emission tomography (PET), and magnetic resonance imaging (MRI).^1,6^ To further measure the intracoronary hemodynamic impact of the anomalous origin, invasive diastolic pressure indices such as the instantaneous wave-free ratio (iFR) can be measured. Testing strategies vary across centers and differ according to coronary morphology.^17,18^ Furthermore, comparing the sensitivity and specificity of testing modalities is challenging in the absence of a universally accepted gold standard. We sought to characterize how coronary morphology is associated with having undergone stress testing, and the odds of ischemia on stress testing.

## Methods

### Patient Selection and Characterization

In this retrospective cohort study, we queried the Cleveland Clinic medical records and billing databases for adult patient encounters (July 2017-April 2023), with diagnosis codes including “anomalous aortic origin of a coronary artery”, “AAOCA”, or “coronary artery malformation” (Figure 1). This search identified 1,303 adults, whose records we reviewed to confirm the AAOCA diagnosis. We excluded other types of coronary artery anomalies (such as coronary artery fistula and myocardial bridges with normal coronary origins). Additional exclusions included hemodynamically significant congenital heart disease (that would independently require intervention) and duplicate patients. After applying these exclusions, 548 adults diagnosed with AAOCA remained. When a patient has multiple anomalous coronaries, one of the anomalous coronaries can have a presumed high-risk feature while the other does not. Therefore, we omitted 17 patients with multiple anomalous coronaries from our analysis cohort, yielding 531 patients with a single anomalous coronary. All patient data was stored in REDCap, with patient consent waived, as approved by the institutional review board (#23 - 146).

**Figure 1:**
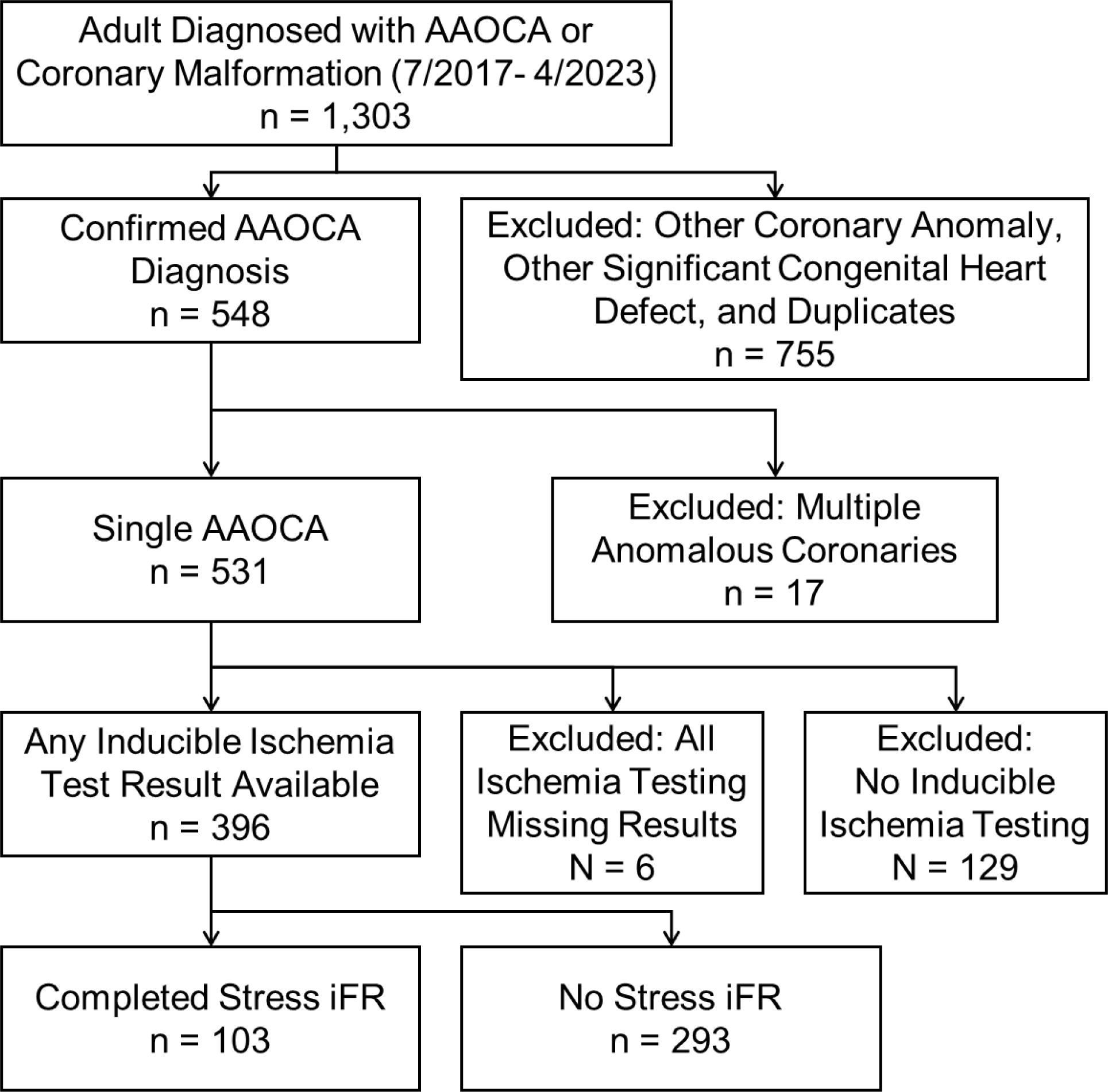
Cohort flow diagram of adults with anomalous aortic origin of a coronary artery (AAOCA) with stress testing for inducible ischemia and sub-cohort with dobutamine stress instantaneous wave-free ratio (iFR) study.

Comorbidities were considered present if documented or if the patient underwent medical therapy for it. Cardiac valvular dysfunction included any grade from mild to severe. Obstructive CAD was defined by stenosis ≥50% in the left main or ≥70% in any of the other major coronary arteries. Arrhythmia included atrial and ventricular types, but excluded aborted sudden cardiac death, which was captured as a separate variable.

### Coronary Morphology Characterization

The coronary morphology for each patient was detailed based on imaging and operative reports. When anatomic information was discrepant, we used a hierarchy of truth, starting with operative reports, computed tomography angiography, MRI, invasive coronary angiography, echocardiography, and clinical notes. We also reviewed the available source images when anatomic details were missing.

Key anatomic features were defined according to the Congenital Heart Surgeons’ Society AAOCA nomenclature. The morphology was characterized by which of the major coronary arteries were anomalous: the right (RCA), left main (LMCA), left anterior descending (LAD), and left circumflex (LCx) coronary. We also determined where it arose from (opposite coronary artery, opposite sinus, noncoronary sinus, or above the sinotubular junction (STJ), and the course type (intramural, interarterial, transseptal, prepulmonic, or retroaortic). Finally, we also determined the presence of other potential high-risk features, such as an acute takeoff angulation (<30°), and a slit-like orifice (with lumen major-minor axis length ratio ≥ 2). Obstructive CAD was defined as stenosis ≥ 50% in the LMCA, or ≥ 70% in the other coronaries. We separately analyzed obstructive CAD in *any* coronary, and in the anomalous coronary. Downstream obstructive CAD in LAD or LCx counted toward obstructive CAD in the anomalous LMCA when it coincided with the territory of ischemia.

### Stress Test Evaluations for Inducible Ischemia

We reviewed all available exercise and pharmacologic stress tests for the 531 patients with a single AAOCA. Stress tests included ECG, echocardiography, SPECT, PET, MRI, and iFR. 396 of the 531 patients (75%) had completed a total of 1,421 individual tests, before any anomalous coronary intervention. Stress test results were coded as normal, indeterminate, and positive for ischemia due to the anomalous coronary artery. ECGs were considered positive if ST elevation was ≥ 1mm in the territory of the anomalous coronary artery.^19^ ST depressions were positive if they were at least 2.0 mm when upsloping, or at least 1.0 mm when horizontal or downsloping at 60-80 ms after the J point. Ventricular arrhythmias and high-grade heart block that worsened with exercise were also positive for ischemia. Baseline bundle branch block and repolarization abnormalities were considered indeterminate. Imaging tests were positive if there was a wall motion abnormality or inducible perfusion defect in the territory of the anomalous coronary. Stress iFR < 0.86 was used as a threshold for ischemia.^1^ Ischemia outside the territory of the anomalous coronary artery was categorized as indeterminate. Absence of ischemia at heart rates less than 85% maximum predicted was also counted as indeterminate.

### Statistical Analysis

Analyses were completed in SAS v9.4 and R v4.5.0. Descriptive statistics included frequency counts for categorical data, means with standard deviations for normally distributed data, and medians with [15^th^, 85^th^] percentiles for skewed data. Patient characteristics considered included age at test, gender, comorbidities, and the anomalous coronary morphology. Due to the strong collinearity between anomalous coronary course types (Table S1), we categorized patients into mutually exclusive groups: transseptal, intramural (without transseptal), interarterial (without transseptal or intramural), and other (prepulmonic or retroaortic). To account for possible association of repeated testing within a patient, a mixed-effect logistic regression (PROC NLMIXED in SAS) with a random intercept was employed. The mixed-effect logistic regression focused on how the patient characteristics and the stress test modality were associated with odds of a positive stress test. Bootstrap aggregation (“bagging”) methodology was implemented to select reliable predictors from a larger list of patient and morphology variables (Table S2).^20^ Forward stepwise selection of 1,000 resamplings with entry and stay criteria of 0.10 and 0.05, respectively, was used. Variables that occurred in ≥50% of the bootstrapped models were considered reliable variables. Based on a priori hypotheses, we included course type, imaging modality, age, gender, and coronary morphology variables in the final mixed-effect model, along with reliable variables. Mean imputation was used for missing values because they were highly populated.

For the sub-cohort with dobutamine iFR results (n=103), we used a random forest regression (randomForestsSRC in R)^21^ to identify patient and morphology variables (Table S2) associated with continuous response-stress iFR. Random forests is a nonparametric statistical ensemble method that utilizes all the variables and makes no distributional or functional (linear or nonlinear) or interaction effects assumptions about covariate relationships to the response.^22^ On-the-fly imputation was used to impute the missing data.^23^ The random forest variable importance (VIMP) measure was used to hierarchically order the variables in relation to the predicted response.^24^ One of the main objectives of the random forest analysis is to be able to visualize, without model assumptions, the relation of features to the response variable. The forest predicts a risk-adjusted relation of selected variable(s) with respect to the response. The result is what is known as “partial dependency plots.” Each partial plot describes the risk-adjusted relationship between the covariate of interest and the response by averaging out the effect of all other covariates in the random forrest model.^25^

## Results

### Patient Characteristics with, Versus Without, Stress Testing

Demographics and comorbidities were broadly similar between patients who underwent ischemia stress testing (n = 396) and those who did not (n = 135) (Table S3). Patients with stress tests were diagnosed more recently, with a median of 6.1 versus 7.5 years (p < 0.001) between diagnosis and cohort cutoff dates (April 2023). When attributes were statistically different, we report values for both groups; otherwise, we highlight values only for the stress-tested group (which formed the cohort of the subsequent logistic regression). Within the stress test cohort, the age at diagnosis was 50.5 ± 16.9 years, and 166 (42%) were female. Cardiac valve dysfunction was present in 129 (33%), hypertension in 247 (62%), hyperlipidemia in 239 (60%), and arrhythmia in 171 (43%). Myocardial bridge over *any* coronary was found in 49 (12%) and over the *anomalous* coronary in 13 (3.3%). When CAD severity was fully quantified, obstructive CAD in *any* coronary was present in 40/388 (11%). Obstructive CAD was present in the *anomalous* coronary in 17/396 (4.7%).

Symptoms at the time of AAOCA diagnosis had some differences in those with versus without stress testing. Chest pain was more common at 266 (67%) versus 72 (53%), p = 0.004. Exertional chest pain was explicitly specified in 144 (36%) versus 32 (25%), p = 0.018. While syncope was present in 69 (17%) versus 8 (5.9%), p = 0.001, no differences were seen for exertional syncope, which occurred in only 8 (2.0%) versus 4 (3.0%), p = 0.52. The remaining symptoms were similar. Among those with stress testing, dyspnea occurred in 234 (59%), myocardial infarction occurred in 36 (9.1%), and aborted sudden death was rare in 8 (2.0%).

Morphologic features of the anomalous coronary were also similar between groups. Among those with stress tests, the anomalous origin most commonly affected the RCA in 262 (66%), followed by the LMCA in 77 (19%), the LCx in 46 (12%), and the LAD in 11 (2.8%). The anomalous coronary arose from the opposite sinus in 299 (76%), opposite coronary in 37 (9.3%), above the STJ in 56 (14%), and noncoronary sinus in 3 (0.8%). The course type was intramural in 186 (47%), interarterial only in 74 (19%), transseptal in 41 (11%), and other in 94 (24%). The length of the intramural course was 9.2 ± 3.8mm. When the coronary dominance was known, the anomalous coronary was more commonly dominant in 215/378 (57%) of those with stress testing, than in 53/119 (45%) of those without it, p=0.036.

### Additional Invasive versus Only Noninvasive Stress Testing Comparison

Differences between patients who underwent invasive dobutamine iFR testing (n = 103) and those with only noninvasive stress testing (n = 293) were more prominent (Table S4). Those with iFR testing were diagnosed younger at 47 ± 15, versus 51 ± 17 years (p = 0.01). The AAOCA diagnosis was also more recent, with only a median of 3.2 versus 7.0 years between the diagnosis and study cohort cutoff dates. Chest pain was similarly common at 70% versus 66%, but the iFR group had less typical angina at 28% versus 39%. Palpitations were more frequent among those with iFR testing, at 27% versus 14%, p = 0.002. Other symptoms were similar, including syncope, dyspnea, aborted sudden death, and myocardial infarction.

Among those with iFR testing, many comorbidities were less frequent than those with only noninvasive stress testing. The less common comorbidities included valve dysfunction at 18% versus 38% (p < 0.001), arrhythmia at 31% versus 47% (p = 0.004), congestive heart failure at 6% versus 20% (p = 0.002), diabetes at 11% versus 24%, and tobacco use at 17% versus 32%. Obstructive CAD in the anomalous coronary was present in only 1.1 % versus 5.5% (p = 0.051). Myocardial bridge over the anomalous coronary was more common in the iFR group at 10% versus 1% (p < 0.001). Other comorbidities, including hypertension and hyperlipidemia, were relatively similar.

Morphologic features were also different for the iFR subgroup, including which coronary was anomalous (p < 0.001) and the course type (p < 0.001). Among those with iFR testing, the anomalous coronaries consisted of 70% RCA, 25% LMCA, 4.9% LAD, and no LCx; the course types were 66% intramural, 19% transeptal, and 13% interarterial only, and 2% other. In contrast, those with only noninvasive stress testing, the anomalous coronary consisted of 65% RCA, 17% LMCA, 2% LAD, and 16% LCx; the course types were 40% intramural, 8% transeptal, 21% interarterial only, and 31% other. The proportions of anomalous coronaries that supplied the posterior descending coronary artery were similar.

### Stress Testing Modalities

The most frequent stress test modality was ECG, with results available in 365 (92%) of all 396 patient with stress testing. The next most common stress test modalities were SPECT in 190 (48%), echocardiograms in 148 (37%), and PET in 126 (32%). Following 2016, there was a gradual decrease in stress echocardiogram and SPECT testing frequency (Figure S2). Stress iFR became available at our center in 2017 and was completed in 103 (26%) patients. MRI was rare, with only 8 (2.0%) patients. Stress tests typically started with ECG, echocardiography, or SPECT, often before AAOCA was diagnosed, and then followed up with PET and iFR (Table S5). The median years from AAOCA diagnosis to the modality’s first stress test was −0.01 [−4.7, 0.95] for ECG, −0.01 [−5.57, 1.75] for echocardiography, −0.04 [−5.24, 2.99] for SPECT, 0.22 [0.03, 1.55] for PET and 0.36 [0.09, 1.48] for iFR, (p < 0.001).

Stress tests were mostly negative for ischemia and often repeated (Table S6). Out of all individual stress ECG tests, only 52/699 (7.4%) were positive for ischemia. Stress echocardiography had a similarly low rate of positive results at 14/198 (7.1%). Among stress SPECT and PET tests, 38/289 (13%) and 44/132 (33%) were positive, respectively. Slightly more than half of the iFR tests were positive at 59/103 (57%).

### Mixed-Effect Logistic Model of Positive Stress Test

In addition to preselected variables (anomalous coronary, anomalous course type, and gender), variables that met statistical threshold in the bootstrap aggregation variable selection for the final mixed-effect logistic regression included age, obstructive CAD, valvular dysfunction, and arrhythmias (Figure 2, Tables S7 and S8). Older age was associated with increased odds of ischemia (p= 0.005). Relative to the average age (50.5 yr), a standard deviation younger (33.6 yr) has a lower odds ratio (OR) of 0.82, and a standard deviation older (67.4 yr) has a higher OR of 1.1. Comparing which coronary was anomalous, only the OR between LMCA versus RCA was borderline significant (OR: 2.1, p = 0.054). The presence of an intramural course also trended toward increased odds compared to prepulmonic and retroaortic courses (OR: 1.8, p = 0.14).

**Figure 2:**
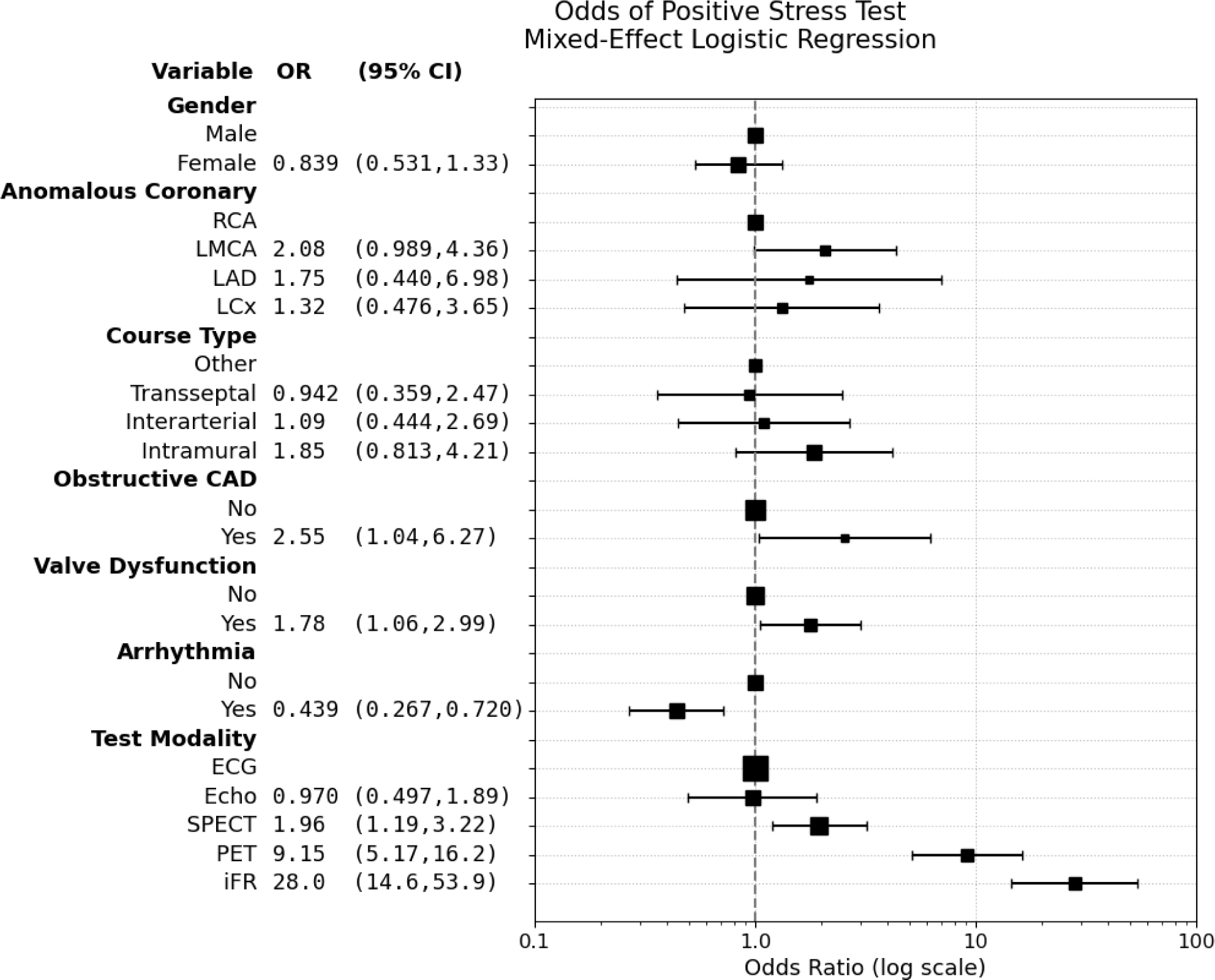
Forest plot of categorical variables in mixed-effect logistic regression. Adjusted odds ratio (OR) with 95% confidence interval (CI) quantified the relative odds of a test showing ischemia in the territory that includes the anomalous coronary artery: right (RCA), left main (LMCA), left anterior descending (LAD), and left circumflex (LCx). Area of the center boxes represents the relative sample size of each group. Obstructive coronary artery disease (CAD) only in the anomalous coronary artery was modeled. Continuous variables are not shown and include older age, which was associated with higher odds of a positive test.

Among comorbidities considered in bootstrap aggregation variable selection, only obstructive CAD, cardiac valve dysfunction, and arrhythmia were statistically significant and reliable. Odds of ischemia increased with obstructive CAD (OR: 2.6, p = 0.042), and valvar dysfunction (OR: 1.8, p = 0.029). Conversely, arrhythmias decreased the odds of ischemia on stress testing (OR: 0.43, p = 0.001).

The strongest predictor for ischemia was the stress test modality. Compared to ECG, iFR had the highest odds (OR: 28, p < 0.001), followed by PET (OR: 9.1, p < 0.001), and SPECT (OR: 2.0, p = 0.008). Echocardiograms had near equal odds of ischemia (OR: 0.97, p = 0.93).

### Random Forrest Model of Dobutamine-Stress iFR

Based on the VIMP, the strongest predictors of stress iFR were the anomalous coronary type and the course (Figures 3 and S1). We also evaluated the effect size by comparing the risk-adjusted median stress iFRs in the partial plots. The anomalous LAD has the lowest risk-adjusted stress iFR (0.75) compared to the anomalous LMCA (0.83) and anomalous RCA (0.84). The course type with the lowest stress iFR was transseptal (0.77), followed by intramural (0.83), interarterial (0.88), and other (0.87). The course type “other” included 2 patients, in whom a prepulmonic LMCA had a stress iFR of 0.90, and a retroarotic LMCA had a stress iFR of 0.91. Anomalous coronaries arising from the opposite coronary or opposite sinus also tended to have a lower iFR (0.80), than for an origin above the STJ (0.87). Notably, these morphologic features associated with lower stress iFR were highly colinear: anomalous LAD, origin from the opposite coronary, and transseptal course (Table S1).

**Figure 3:**
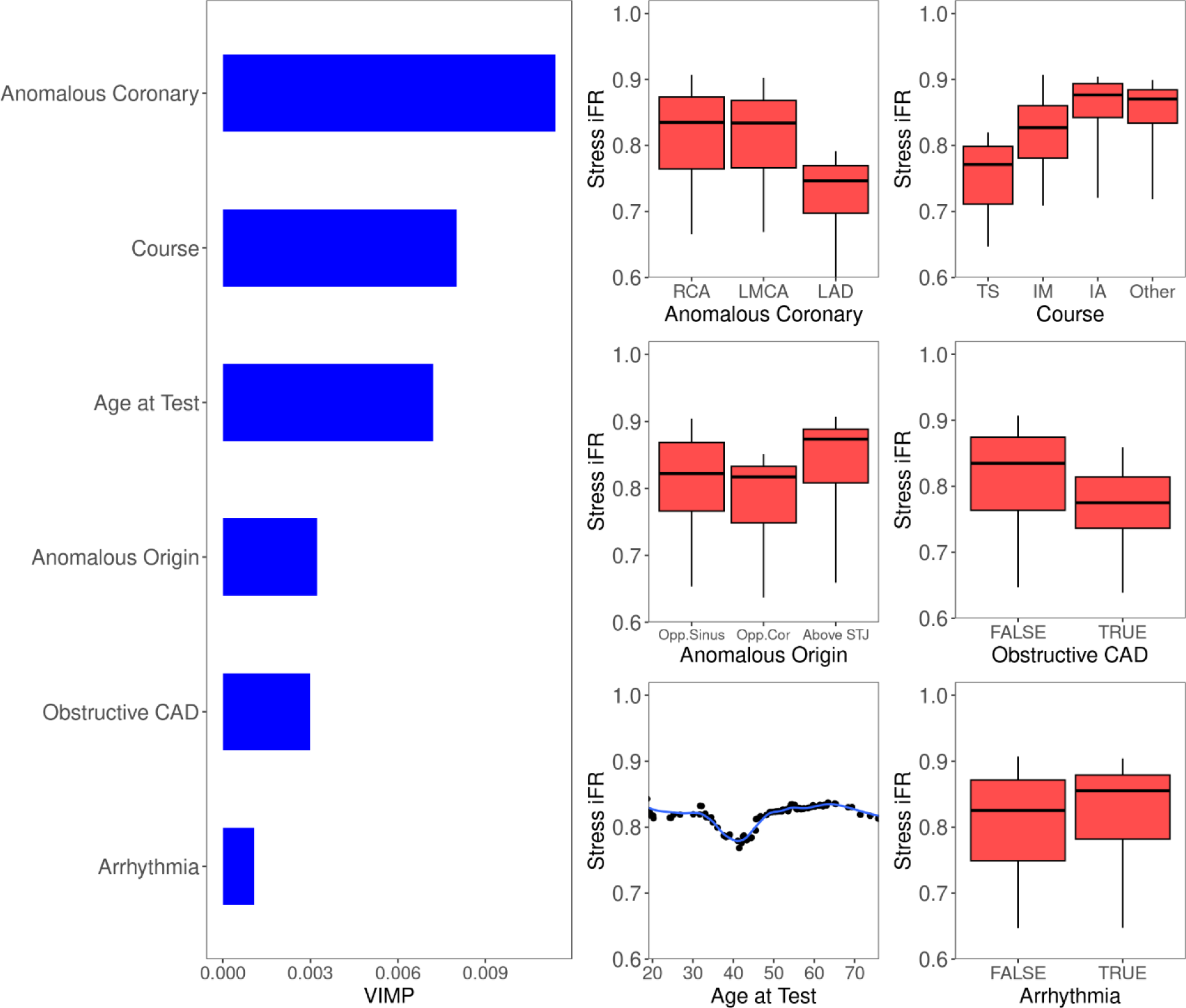
Random forest analysis of dobutamine-stress iFR with predictors including patient characteristics and coronary morphology. Variable importance (VIMP) and partial plots of stress iFR for the top 6 variables are shown. Mutually exclusive course type included transeptal (TS), intramural (IM), interarterial (IA), and other (prepulmonic and retroaortic). Abbreviations included opposite (Opp.).

Among comorbidities assessed, obstructive CAD lowered the stress iFR from 0.84 to 0.78. Other predictor variables had low VIMP and a small effect size. Age had a non-monotonic relationship with stress iFR, ranging from 0.80 to 0.84. History of arrhythmias was associated with a higher stress iFR, increasing from a median of 0.83 to 0.86. Cardiac valve dysfunction had a slightly negative VIMP and minimal effect on the iFR, decreasing the median iFR from 0.84 to 0.82.

**Table 1:**
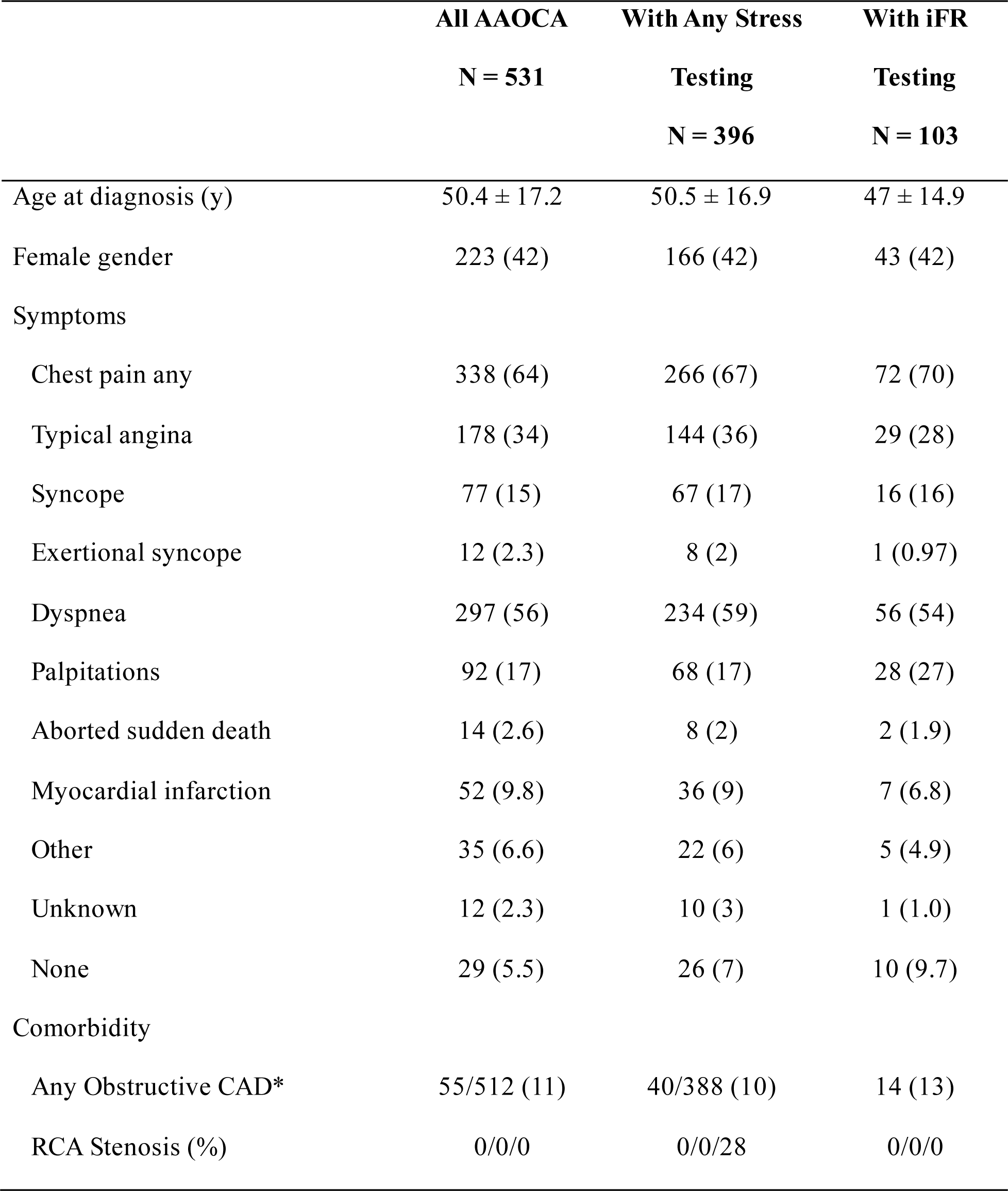
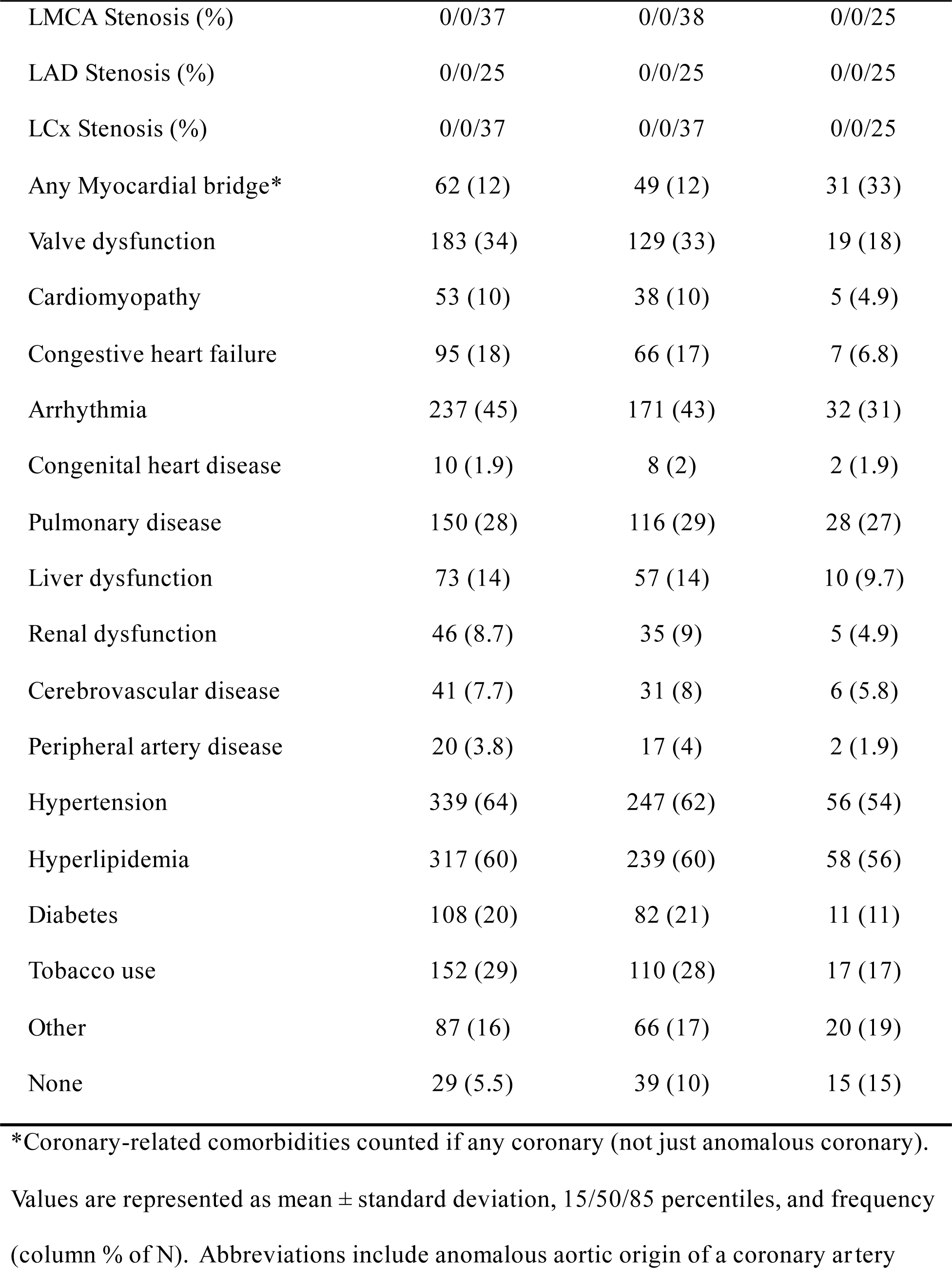
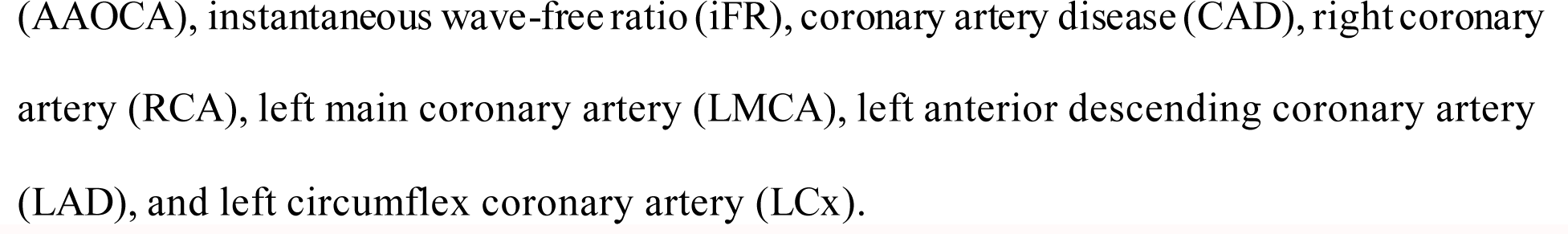
Patient characteristics for adults with an anomalous aortic origin of a coronary artery and sub-cohorts with specific stress test results.

**Table 2:**
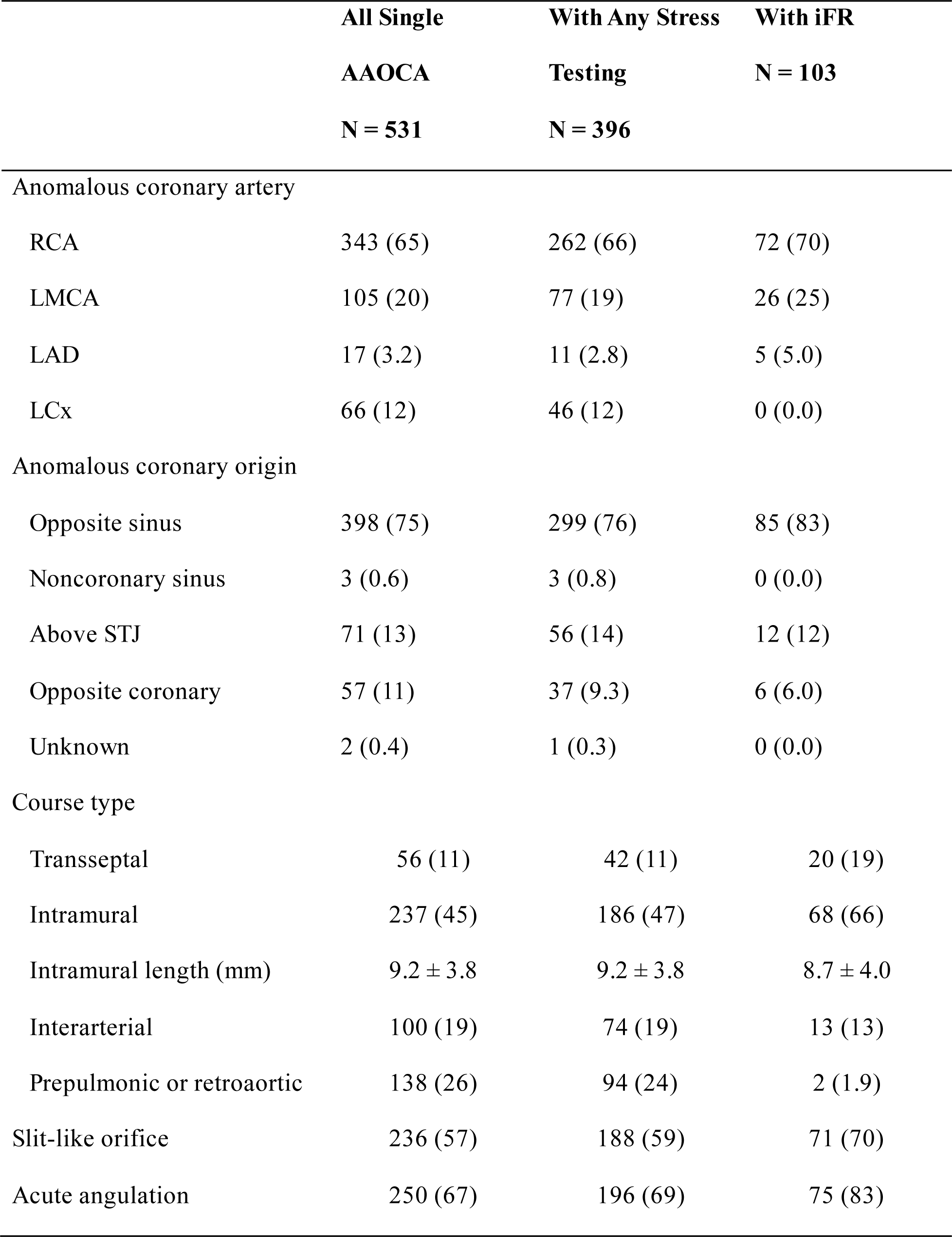
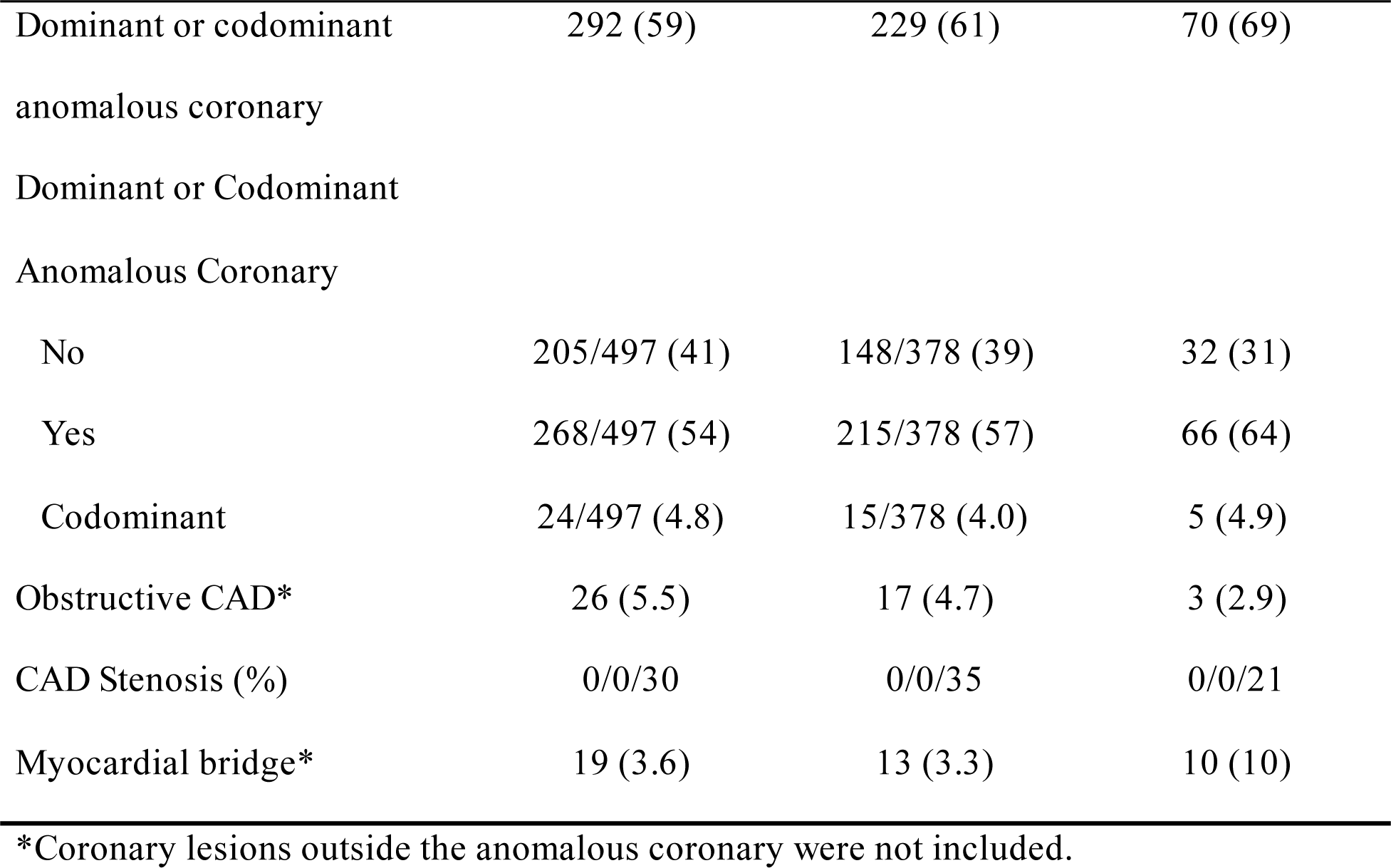
Anomalous coronary anatomy and coronary comorbidities in the anomalous coronary.

## Discussion

Ischemia evaluation with stress testing in AAOCA is often needed to guide surgical repair, especially when the cause of potential cardiac symptoms is unclear.^1,12^ However, the frequency and test modalities vary widely across centers, and it remains unclear which modality best detects ischemia.^6^ This single-center study provides insights into key questions: (1) whether patients with presumed higher-risk coronary morphology features were more likely to undergo stress testing, (2) which of these features increased the odds of a positive test, and (3) how the test modality influenced the odds of detecting ischemia. To our knowledge, this is the largest adult AAOCA study correlating coronary morphology to ischemia stress test results, and the only one to integrate the relative odds of ischemia across multiple test modalities.

### Stress Testing Practice Patterns

The proportion of AAOCA patients who underwent stress testing (75%) was relatively high compared to other AAOCA registries.^6,9,18,26^ Therefore, the potential impact of missing stress test results from incomplete medical records is small. Some patients may have undergone surgical repair without stress testing, because current guidelines state that surgical repair is reasonable for high-risk AAOCA morphologies (even without symptoms or a positive stress test).^12,27^ However, our congenital coronary anomalies program agrees with the recommendation by the Congenital Heart Surgeons’ Society to obtain baseline stress testing for risk stratification and postoperative comparison.^6^ Meanwhile, some clinicians may have been under the impression that AAOCA poses minimal risk in older adults, reducing the need for stress testing. More recent publications address that AAOCA carries cardiovascular risk in older adults as well, with the potential need for surgical repair.^12,14,16^ This increasing awareness may explain why those who underwent stress testing were diagnosed in a more recent era by 1.4 years.

We initially hypothesized that patients with high-risk coronary morphology would be associated with any stress testing, either via increased cardiac symptoms or clinical concern for inducible ischemia based on the anomalous coronary artery. The increased prevalence of chest pain, dyspnea, and syncope, in the stress-tested group, reflects concern for myocardial ischemia from AAAOCA and or CAD in this cohort with an average age >50 years.^14^ While symptoms were associated with stress testing, the presence of an anomalous left LMCA (versus RCA), intramural course, slit-like orifice, or acute angulation was not. These morphologic features are well recognized as ischemia and sudden cardiac arrest risk factors among children and young adults.^2,6^ In our study, only having an anomalous coronary as the dominant coronary was associated with more stress testing. Anomalous origin of the dominant coronary could potentially increase the probability of ischemia and prompt stress testing. The lack of a clear association, between coronary morphology and stress test completion, is partly due to many patients having undergone stress testing before their AAOCA diagnosis. Consequently, many low-risk AAOCA variants, such as the anomalous LCx with a retroaortic course, also had some stress testing as part of the initial evaluation for possible CAD. Ultimately, the similarity of morphology and comorbidities with stress test completion suggests that logistic regression results are generalizable to the broad spectrum of AAOCA morphologic variants at our center.

While the initial stress ECG, echocardiography, and SPECT tests commonly preceded the AAOCA diagnosis, stress PET was almost always performed after AAOCA diagnosis. Even if perfusion imaging with SPECT was already completed, dobutamine PET was still obtained. Dobutamine PET better mimics exercise physiology and detects ischemia with superior sensitivity than SPECT which typically used adenosine or regadenoson to induce hyperemia.^28,29^ Dobutamine PET was also more readily available at our center than stress MRI, which was rarely completed in this AAOCA cohort.

Invasive dobutamine iFR was reserved for higher-risk AAOCA variants, or those requiring further risk stratification. It also never included the anomalous LCx, which was almost always retroaortic without an intramural course and considered benign. Invasive iFR testing was also helpful for evaluating concomitant myocardial bridge or CAD to determine the exact cause of ischemia. The higher prevalence of myocardial bridges in the iFR cohort is similar to the published prevalence of about one-third in the general population, likely reflecting better detection by angiography.^30^ The less frequent cardiovascular comorbidities, such as obstructive coronary artery disease and valve dysfunction, within the iFR sub-cohort may be a result of selection bias and with younger patients.

### Impact of Anomalous Coronary Morphology on Ischemia

The mixed-effect logistic regression results are consistent with previous findings that the anomalous LMCA and intramural course trended toward increased risk of ischemia.^6,31^ The higher risk of ischemia with the anomalous LMCA may be due to the relatively greater blood flow, supplying the larger territory of myocardium. With an OR of 2.1, the effect size of the anomalous LMCA was only slightly smaller than the OR of 2.6 for obstructive CAD, a widely accepted indication for coronary intervention.^32^ Course type, on the other hand, only trended toward statistical significance. Nonetheless, with an OR of 1.9, the presence of an intramural course remains clinically significant to assess, especially with the evidence reported in other registries.^6,31^

The Congenital Heart Surgeons’ Society AAOCA registry, (with patients diagnosed before age 30), found ischemia to be more common for the anomalous LMCA, intramural course, longer intramural course, high takeoff, or slit-like ostium.^6^ The older patients in this study have higher rates of comorbidities, such as CAD, that may outweigh the ischemic impact of the anomalous origin. The rising vascular stiffness with age could also attenuate the dynamic compression of the intramural course during exercise. A recent large single-center report found no association between morphology and stress test results, but the sample size was smaller, with only 62/173 (36%) having undergone any stress testing.^9^ The study by Stark et al. had a similar middle-aged adult cohort and found no association between the intramural length and ischemia.^31^ Rather, they found a strong association between dobutamine fractional flow reserve and the intramural minimal luminal area, a measurement we did not collect in this study. Patient-specific 3D-printed and *in silico* models have also demonstrated how the intramural course can be restrictive and become narrower during exercise, potentially leading to ischemia.^7,33,34^

The dobutamine-stress iFR subanalysis provided additional insight into the hemodynamic impact of the transseptal LAD. The strong collinearity between the anomalous LAD, transeptal course, and origin from the opposite coronary makes it difficult to parse the relative contribution of each factor toward the ischemia burden. Furthermore, interpretation of these findings must be tempered by the possibility of selection bias. Since the transseptal course was historically regarded as benign,^13^ perhaps only the most symptomatic cases of transseptal AAOCA were referred for iFR evaluation. The recent development of the Najm transconal unroofing provides a presumably more robust surgical treatment than traditional bypass grafting, and has increased referrals.^35^ The relatively high rates of inducible ischemia from the transseptal course is also corroborated by a report by Doan et al., showing that half of transseptal courses in children have positive dobutamine-stress MRI and invasive coronary pressure ratio indices.^36^ Whether lower iFR in the transeptal LAD reflects true ischemic burden or a measurement bias warrants further investigation.

### Stress Test Modality Performance in AAOCA

Because there is no definitive standard for diagnosing ischemia in AAOCA, it is difficult to determine the sensitivity and specificity of each stress test modality.^6^ Repeated testing also increases the likelihood of false positives. To address these challenges, we applied a mixed-effect model to estimate the relative odds of detecting ischemia while minimizing bias. After adjusting for covariates, test modality was the most robust predictor of a positive result.

Our findings align with the relative stress testing performance for CAD.^37–39^ iFR had the highest odds of detecting ischemia, followed by PET and SPECT. With increased use of PET and iFR in recent years, our center has reduced reliance on stress echo, and SPECT (Figure S2). Notably, our center predominantly utilizes PET instead of MRI for perfusion imaging; both modalities have similar sensitivity and specificity in patients with obstructive CAD. ^40^ Despite our decision to use a more severe threshold of <0.86^1^ (instead of the commonly used ≤0.89 threshold for revascularization in stable CAD^41,42^), we had relatively high odds of detecting ischemia with this modality. Bigler et al. also found dobutamine-stress fractional flow reserve to be 2.7 and 7 times more likely to detect ischemia than PET and SPECT, respectively, for the anomalous RCA.^43^ The low odds of stress echocardiography and ECG for detecting ischemia generally follow the ischemia cascade pathophysiology, where perfusion abnormalities precede wall motion abnormalities on echocardiography, and finally ECG changes.^19^

The relative odds of a positive test in AAOCA are critical for selecting stress test modalities and interpreting the results. When the goal is to minimize the false negative rate, especially for high-risk coronary morphologies, our data favors the utilization of dobutamine-stress iFR for further risk stratification. Importantly, iFR testing is an invasive catheter-based evaluation, which has its own procedural risks that must be weighed into the stress test modality selection. Therefore, a comprehensive evaluation of the ischemia pretest probability remains essential and should include the symptoms and anomalous coronary morphology.

To ultimately determine the best stress testing modality for AAOCA, it needs to be correlated with presenting typical symptoms such as angina and major adverse cardiovascular events, such as sudden cardiac arrest or myocardial infarction. During our retrospective records review, we found it difficult to ascertain the exact nature of chest pain, though it was frequent in 67% of the patients who underwent stress testing. The presence of chest pain with exertion was often inconsistent, and many patients had sedentary lifestyles, limiting our ability to grade the type and severity of angina. Aborted sudden death and myocardial infarction, at 8 (2%) and 36 (9%), respectively, were not frequent enough for robust statistical modeling of the performance of multiple test modalities. The study by Gentile et al., with a similar middle-aged adult cohort, did not find an association between inducible ischemia on testing and cardiac events, potentially due to insufficient sample size and infrequent events.^9^ Thus, a larger cohort and continued follow-up for cardiac events are necessary to evaluate the prognostic implications of ischemia on stress testing.

### Comorbidities Confounding Stress Testing Results

The mixed-effect logistic regression model showed increasing odds of ischemia with age, which correlates with various comorbidities, including CAD, and degenerative valve disease. Obstructive coronary artery disease, present in 5% of the anomalous coronaries, significantly increased the odds of ischemia as expected.^39^ Given the often atypical chest pain and potential increased prevalence of myocardial bridges in some AAOCA cohorts, we increasingly also evaluate for concomitant myocardial bridges.^44^ Valvular dysfunction also increased the odds of ischemia, potentially due to increased myocardial demand.^45^ Our data could not specify what type of valve disease was present, which greatly our ability to interpret its clinical significance. However, if aortic valve stenosis or regurgitation were present, the resulting decreased coronary perfusion pressures could also lead to ischemia.^46^ On the other hand, arrhythmia had the opposite effect. Although we could not subclassify the exact type of arrhythmia, we believe that the vast majority was likely atrial, as we classified aborted sudden death as a separate variable. Patients with tachyarrhythmias may have been on a rate-limiting agent that limited their ability to achieve at least the 85% maximum predicted heart rate threshold. The submaximal test in such patients or baseline chronotropic insufficiency lowers the odds of detecting ischemia.^19^ Thus, comorbidities can contribute to true myocardial ischemia but also obfuscate the degree to which AAOCA is causing ischemia, for which surgery would be indicated.

### Limitations

This retrospective study may be limited by potential unaccounted confounders that may be correlated with stress test selection and performance. Morphologic characterization was largely categorical and may not fully capture the impact of the complex 3-dimensional geometry and vascular stiffness on coronary blood flow. The high degree of collinearity within comorbidities and morphologic features (i.e. intramural course, slit-like orifice, and acute angulation) makes it difficult to statistically determine which of the collinear factors is truly causing ischemia. Finally, referral bias in the single quaternary care center may skew results toward higher-risk and more symptomatic patients, which may limit generalizability to all AAOCA patients.

## Conclusions

In this large retrospective single-center study, stress testing for ischemia was frequently performed in adults with AAOCA. Coronary morphology was similar between patients who underwent stress testing and those who did not. However, those who underwent the invasive dobutamine-stress iFR were more likely to have high-risk features or requiring further risk stratification, such as having an intramural or transseptal course. Similarly, it was never performed in an anomalous LCx that is considered a benign coronary anomaly. Among patients who completed any stress testing, the anomalous LMCAwas strongest *morphologic* predictor of ischemia. The strongest impact on the test result was the modality, with stress PET and iFR more likely to detect ischemia than stress ECG. Effective risk stratification for guiding AAOCA surgical repair requires integrating symptoms, coronary morphology, and stress testing results, while accounting for performance characteristics of each stress test modality. The procedural risks of invasive evaluation must be factored into decision-making. Continued longitudinal follow-up is needed to determine the optimal stress testing strategy.

## Data Availability

Patient data is currently not authorized to be shared.

## Acknowledgements

The Cleveland Clinic Adult Anomalous Aortic Origin of the Aorta Working Group collectively reviewed an entered data utilized in this study: Ellen K. Brinza MD MS, Michael J. Haupt MD, Christiane Mhanna DO, Dominique L. Tucker MD, Sohini Gupta MD, and Miza Salim Hammoud MD MS. We also recognize William G. Williams MD and the Congenital Heart Surgeons’Society Center for Research and Quality for their guidance on variable definition s.

## Abbreviations

AAOCA: anomalous aortic origin of a coronary artery
CAD: coronary artery disease
iFR: instantaneous wave-free ratio
LAD: left anterior descending (coronary artery)
LCx: left circumflex (coronary artery)
LMCA: left main coronary artery
RCA: right coronary artery
STJ: sinotubular junction
VIMP: variable importance

## Supplemental Material

Tables S1-S8

Figures S1-S2

